# Total and regional skeletal muscle mass references by sport participation and body size in youth

**DOI:** 10.1101/2023.02.07.23285593

**Authors:** Lorena Correas-Gómez, José Ramón Alvero-Cruz, Jesús Barrera-Expósito, Margarita Carrillo de Albornoz-Gil, Ana L. Quitério, Elvis A. Carnero

## Abstract

Skeletal muscle mass (SMM) is a relevant indicator of adolescent health influenced by sport participation, body size, and maturation. However, limited data are available from techniques accessible to field professionals.

**Purpose:** To provide reference data of regional and total-body SMM and fat-free mass (FFM) derived from anthropometry among groups of age, sex, and sport participation in healthy Caucasian children and adolescents.

**Materials and methods:** A cross-sectional dataset of 1,438 participants aged 8-18 years were analyzed for this study. Regional and total-body SMM and FFM were estimated from anthropometric variables. Sport participation was obtained by *ad hoc* questionnaires and maturity offset was calculated using predictive equations. General linear model of mixed factors was used to analyze the variance of FFM or SMM across groups and confounders. Ln-ln regression analysis was applied to describe the scaling relationship between SMM and height.

**Results:** Positive interactions were found between sex, sport participation, and age for regional muscle variables (*P*<0.05). Adjusted total and regional SMM significantly increased along school-age periods in the active groups (∼2.2 kg gain, *P*<0.05, boys and girls; arm, ∼1.3 cm gain, *P*<0.01, boys), and allometric values were higher in boys than girls for regional muscle (*P*<0.01).

**Discussion:** The novel contribution of this analysis includes regional indicators of SMM and the relevance of sport participation on SMM accrual in post-pubertal boys. Also, confirms that muscle growth rate varies across sex and phases of puberty, which arises a plausible sexual phenotype/dysmorphism of regional SMM influenced by sport participation.

## INTRODUCTION

Interest in the development and assessment of skeletal muscle mass (SMM) of children and adolescence has increased ^1^ due in part to concerns for the healthy growth and motor development ^2^. Moreover, muscular fitness and metabolic risk in youth are related ^3^, while longitudinal observations indicate a relationship between muscular strength early in life and adult cardiovascular risk ^4^ and the metabolic syndrome ^5^. Additionally, muscle mass and strength are related with skeletal development ^6^ and quality of bone mass ^7^.

Although important, direct measurement of SMM is limited to imaging methods, which are expensive and not widely available ^8^, and at times impractical with children ^1^. Whole body molecular level methods are often used to estimate fat-free mass (FFM), a surrogate for SMM. These methods have provided the basis for the development of reference data based on both laboratory ^9–13^ and field methods, specifically bioelectrical impedance analysis (BIA) ^14,15^ and anthropometry ^16^. Anthropometry is less dependent on environmental error (for example, previous meal, hydration or temperature) than BIA, and provide an estimates of regional ^17^ and total-body muscle mass in youth ^18^ with acceptable validity, although the need to quantify measurement variability is often overlooked.

Regardless of the assessment method, the comparisons between amounts of SMM across sex- and age-groups require accounting for differences in biological maturation and body size. During the adolescent growth period, individuals of the same chronological age may vary considerably in height and maturity status ^19^. Consequently, these confounding factors need also to be controlled when examining the independent effect of physical activity or exercise training, as participation in regular physical activity programs plays a positive role in lean mass accrual ^20,21^.

To our best knowledge, there is a lack in the availability of combined reference data for regional and total SMM derived from field techniques along with the analysis of regular participation in sport, maturation, and body size. The combined reference data may contribute to identify differences in age- and velocity-patterns of muscle distribution at a regional level compared to total-body SMM because lower limbs may develop first compared to upper extremities ^22^. Moreover, the use of an inexpensive and accessible technique like anthropometry would benefit a wide range of field professionals (such as physical education teachers, coaches, nutritionists, and pediatricians) to help to monitor SMM evolution across age. Finally, reporting data separately for children and youth participating in regular sports practice (SP) and those who are not involved in sport participation (NSP), would allow for comparisons of population-specific data and specific regional adaptations as consequence of exercise training.

Thus, the objectives of the current study are to: 1) describe the development of regional and total SMM estimated from anthropometry in children and adolescents of both sexes; 2) analyze the development of SMM across school-age groups of sport participation (SP vs. NSP) and maturity status ; and 3) provide reference data for children and adolescents of European (Caucasian) ancestry.

## MATERIALS AND METHODS

### Study design

The data comprising the base for this study were based on several cross-sectional anthropometric surveys of children and adolescents conducted between 2006 and 2015. The participants were from the Northwestern, Central, Eastern and Southern regions of Spain. Data from an independent study in youth athletes from Portugal were available and polled together for statistical analyses.

The participants were volunteers from previous studies from our labs, who were recruited from local schools and sport clubs through written and oral advertisements. Each participant and one parent were informed about the research procedures; afterwards, written informed consent was signed by parents of children or adolescents who agreed to participate in the study. Inclusion criteria were age between 8-18 years old (yrs), free of acute or chronic diseases, and normal morphology (i.e., no total or partial limb amputations). All procedures and consent forms were approved by the Ethical Committee Board of the Faculty of Medicine of the University of Málaga for studies in Spain.

### Sample

The initial sample included 1,659 children and adolescents aged 8-18 yrs. The analysis was limited to children and adolescents of European (Caucasian) ancestry due the low numbers of other ethnicities, and those with complete anthropometry were retained for analysis. The final sample included 1,438 healthy youth (66.8 % boys and 33.2 % girls, mean age = 13.63 ±2.60 yrs; mean body mass index = 20.22 ±3.50 kg/m^2^), out of which 77.2% were involved in regular sports programs (n = 1081, 25.4% girls, 38 missing).

### Procedures

Body weight was measured with a digital scale (Tanita®, model UM-060) to the nearest 0.1 kg wearing light clothes; height was measured with a portable stadiometer (Tanita® Leicester) to the nearest 0.1 cm. A standard anthropometric procedure was performed to measure skinfolds thickness and circumferences on the right side of the body, which were assessed by a calibrated caliper (Holtain) to the nearest 0.1 mm and a flexible steel tape to the nearest 0.1 cm respectively. Limb circumferences (C_limb_) were measured with participants in a relaxed standing position at three sites following this protocol: upper arm, mid-distance between the lateral and superior part of the acromion border and the proximal and lateral border of the radius head with the arm hanging by the side; thigh, mid-distance between the inguinal crease and the anterior surface of the patella on the anterior midline; and calf, at the maximum circumference. Skinfolds (SKF) were obtained at the same level as the C_limb_, specifically, parallel to the long axis of the posterior upper arm for triceps, of the thigh for anterior thigh, and, of the leg with the foot placed on a box and the knee bent at an approximate 90-degree angle for medial calf. All measurements were obtained by four trained anthropometrists. The technical error of measurements (TEM) for intra-observer (anthropometrist) ranged from 0.39 to 0.47 mm for SKF and from 0.04 to 0.35 cm for C_limb_ measurements. The inter-observer TEM was 0.27-0.47 mm and 0.04-0.46 cm for SKF and C_limb_ respectively (all <2%).

#### Fat-free mass

Fat mass (FM) was predicted using the equations of Slaughter et al. (1988) from sum of SKF triceps and calf ^23^, and in turn FFM was derived by subtracting FM from body mass.

#### Total-body SMM

Two age-specific models were used to estimate SMM (kg) from anthropometric variables ^18,24^. Poortmans’ model ^18^ was used with youth under 16 yrs and Lee’s model ^24^ was used for youth 16 yrs and older following the recommendation suggested by Kim et al. for maturation-related variation in SMM distribution ^25^. The authors found that the relation between total-body SMM and appendicular lean soft tissue (SMM/ALST) was stable in adults but not in children and adolescents, which varied depending on their maturation level, thus, it varied in young at Tanner stage <5 whilst remained stable in the more mature ones (Tanner stage 5). In this study, data relative to Tanner stages was not available in all participants, so we used an alternative age cut-off value, assuming that all children aged 16 and beyond had achieved stage 5 for pubic hair, breast and genital development ^26^.

Lee’s model was developed and cross-validated against MRI (*R*^2^ = 0.91, SEE = 2.2 kg; equation [1]) and the same equation was later used with adapted coefficients for child- and adolescent populations by Poortmans et al. (*R*^2^ = 0.966, SEE not reported; equation [2]). The equations were as follows:

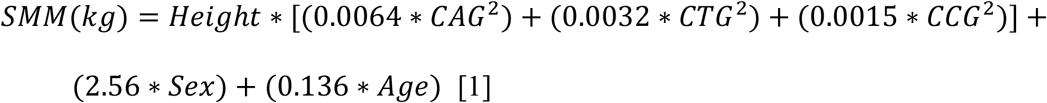

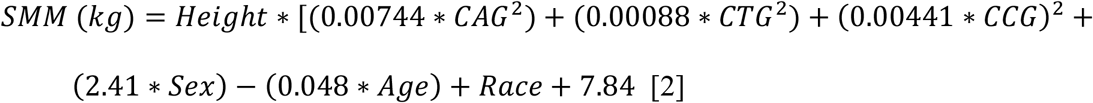

where height was in m, corrected arm girth (CAG), corrected thigh girth (CTG) and corrected calf girths (CCG) were in cm, age in years, sex was 0 for female and 1 for male, and race was 0 for white.

#### Regional FFM

Muscle circumferences (Mc) were estimated from each C_limb_ and its corresponding SKF (equation [3]) which were used to estimate corrected girths at three sites: Mid-upper arm (arm corrected girth, ACG), mid-thigh (thigh corrected girth, TCG), and calf (calf corrected girth, CCG) ^27^.

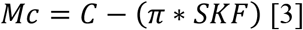

#### Sport Participation

Information about participation in regular sports clubs/teams of each volunteer was obtained by *ad hoc* questionnaires. Participants were classified as sport participants (SP) if they were involved in organized sport practice a minimum of 3 times per week and ≥1-hour practice per workout as extracurricular activities (involving competition or not), and otherwise as non-sport participants (NSP). Organized physical activities like running, dancing and resistance training were considered as SP in this study. The questionnaires were completed by the participants and supervised by the evaluators on site to ensure that every question was completed by each participant. Among those children who could not answer properly or had doubts about their sport practices, we asked for help from their parents, teachers, or coaches.

#### Maturity status

Maturity offset (MO) was predicted from age and height in boys (*R*^2^ = 0.896; SEE = 0.542) and girls (*R*^2^ = 0.898; SEE = 0.528) ^28^. Subjects with a negative predicted MO were classified as pre-peak height velocity (PHV), while those with a predicted MO of zero or higher were classified as post-PHV. Additional maturational groups were defined by age ranges according to phases of puberty (pre-puberty, puberty, and post-puberty) as follows: 8-10, 11-13, 14+ yrs for girls, and 8-12, 12-15, 16+ yrs for boys.

### Statistical Analyses

Subjects were categorized into single chronological age (CA) groups based on the whole year (i.e., 8 yrs = 8.00 to 8.99) and by sex. Children and adolescents were also classified as thin, normal weight, overweight or obese using the BMI and sex-specific criteria of the International Obesity Task Force (IOTF) ^29^.

For further analysis, participants were classified according to school-age periods in Spain: <12 yrs in Primary School (P-S), 12-15 yrs in Secondary School (S-S) and >15 yrs in High School (H-S). This classification is in accordance with changes in PE curriculum from one period to another. An overview of the evolution of differences in SMM between groups along school-age periods could be interesting for PE teaches, coaches, and pediatricians. Estimated means of SMM (adjusted by age and height) for each group at every period can be found in the supplementary material.

Descriptive data are expressed as means and standard deviation or median and ranges when variables did not have normal distribution as determined by Shapiro-Wilk test. FFM and SMM were expressed as absolute (kg) or relative to body weight (%). Simple regression analysis was used to explore the relationship between main variables with age and height, and individual slopes were compared for sex and SP/NSP groups. General linear model of mixed factors was used to analyze variance of FFM or SMM variables among age and school-age groups, and interactions: age x sex, age x sport participation, age x sex x sport participation, school-age periods x sex x sport participation. Tukey’s multiple comparison tests were carried out to detect which groups differed from each other. Additionally, a simple allometric model was applied in natural logarithmic (ln) form to describe regional and total muscle mass scaling relations to height ^30^.

A type II error of 0.05 was considered for all tests (two tailed) and effect size and 95 % confidence intervals (CI) were reported. Statistical procedures were performed using SPSS® statistics software (version 15.0; IBM Corporation, Armonk, NY, USA) and GraphPad Prism (version 7; GraphPad Software, La Jolla, CA, USA).

## RESULTS

The sample included 74.4 % normal weight (732 boys, 337 girls), 16.2 % overweight (147 boys, 85 girls) and 3.5 % obese (30 boys, 20 girls) participants. Thinness had a prevalence of 5.9 % (51 boys, 34 girls). Sample characteristics by sex and CA categories indicated that in girls, median of weight and height increased until ages of 15 and 17 respectively, whereas in boys it increased up to 18 yrs. Sex differences were not significant before age of 14, except at 11 and 13 yrs when girls were heavier and shorter than boys respectively (*P*<0.05, table I).

**Table I.** Characteristics of the sample by age and sex groups.

| **Group** | **N** | **Sex** | **Age (yrs)** | **Weight (kg)** | **Height (cm)** | **BMI (kg/m<sup>2</sup>)** | **BF (%)** |
| --- | --- | --- | --- | --- | --- | --- | --- |
| 8 | 25 | Girls | 8.3 ± 0.3 | 30.3 ± 6.9 | 131.4 ± 4.6 | 17.4 ± 3.3 | 22.5 ± 7.4 |
|  | 46 | Boys | 8.5 ± 0.3 | 29.1 ± 5.1 | 130.6 ± 6.2 | 17.0 ± 2.3 | 14.3 ± 6.3 |
| 9 | 31 | Girls | 9.5 ± 0.4 | 34.8 ± 5.3 | 136.6 ± 5.7 | 18.6 ± 2.5 | 26.2 ± 6.1 |
|  | 54 | Boys | 9.5 ± 0.3 | 34.2 ± 7.2 | 137.1 ± 5.7 | 18.1 ± 2.8 | 17.1 ± 7.5 |
| 10 | 24 | Girls | 10.2 ± 0.4 | 37.1 ± 9.8 | 143.2 ± 9.0 | 17.9 ± 2.7 | 22.6 ± 5.6 |
|  | 65 | Boys | 10.5 ± 0.3 | 36.6 ± 7.2 | 141.8 ± 6.7 | 18.1 ± 2.7 | 19.6 ± 7.2 |
| 11 | 45 | Girls | 11.4 ± 0.3 | 44.4 ± 11.1 | 149.1 ± 8.5 | 19.8 ± 3.4 | 25.7 ± 6.7 |
|  | 100 | Boys | 11.5 ± 0.3 | 39.8 ± 6.7 | 147.1 ± 6.8 | 18.3 ± 2.3 | 19.4 ± 6.8 |
| 12 | 38 | Girls | 12.5 ± 0.4 | 46.4 ± 11.5 | 153.0 ± 7.1 | 19.7 ± 3.5 | 23.8 ± 8.3 |
|  | 114 | Boys | 12.4 ± 0.3 | 46.0 ± 10.5 | 153.8 ± 8.2 | 19.3 ± 3.1 | 20.5 ± 8.6 |
| 13 | 73 | Girls | 13.4 ± 0.3 | 50.6 ± 9.6 | 158.4 ± 7.3 | 20.1 ± 3.1 | 23.4 ± 5.9 |
|  | 108 | Boys | 13.4 ± 0.3 | 50.8 ± 11.2 | 161.3 ± 8.5 | 19.4 ± 3.1 | 17.8 ± 8.9 |
| 14 | 73 | Girls | 14.4 ± 0.3 | 53.4 ± 10.0 | 159.9 ± 5.9 | 20.8 ± 3.5 | 25.7 ± 7.3 |
|  | 119 | Boys | 14.4 ± 0.3 | 58.3 ± 14.6 | 166.8 ± 8.9 | 20.8 ± 4.1 | 17.8 ± 10.2 |
| 15 | 67 | Girls | 15.4 ± 0.3 | 58.2 ± 9.7 | 162.0 ± 7.4 | 22.2 ± 3.8 | 28.1 ± 7.8 |
|  | 131 | Boys | 15.3 ± 0.3 | 66.5 ± 12.0 | 174.2 ± 9.7 | 21.8 ± 3.2 | 17.4 ± 7.9 |
| 16 | 52 | Girls | 16.4 ± 0.3 | 56.7 ± 11.5 | 162.8 ± 7.9 | 21.3 ± 3.5 | 26.2 ± 7.7 |
|  | 116 | Boys | 16.3 ± 0.3 | 67.4 ± 12.2 | 175.1 ± 8.6 | 21.9 ± 3.1 | 15.6 ± 7.5 |
| 17 | 35 | Girls | 17.3 ± 0.3 | 59.1 ± 7.5 | 165.9 ± 8.3 | 21.6 ± 2.0 | 25.7 ± 6.4 |
|  | 72 | Boys | 17.3 ± 0.3 | 70.6 ± 11.7 | 177.0 ± 10.6 | 22.5 ± 2.7 | 15.1 ± 6.2 |
| 18 | 15 | Girls | 18.1 ± 0.2 | 57.5 ± 7.8 | 164.5 ± 7.0 | 21.2 ± 2.4 | 25.1 ± 6.9 |
|  | 35 | Boys | 18.2 ± 0.3 | 71.5 ± 7.9 | 177.7 ± 7.6 | 22.6 ± 2.0 | 12.4 ± 4.6 |

Group, subjects grouped annually by years of age; BMI, body mass index; BF, body fat.

Relative SMM increased with age and reached the highest value at 18 yrs in boys and at 17-18 yrs in girls (table II). In all age groups, boys had greater mean values of %SMM and %FFM than girls except for %FFM at 10 yrs (all *P*<0.01). Additionally, the spurt of SMM (kg/year) occurred one year earlier in boys (3.80 ±0.42 kg/year gain from 14 to 15 yrs) than in girls (2.6 ±0.48 kg/year gain from 15 to 16 yrs; both *P*<0.000), but at similar ages if expressed as % of body weight (from 15 to 16 yrs, 3.53 ±0.45 %/year in boys and 5.39 ±0.58 %/year in girls; both *P*<0.000). Regional muscle variables are presented as Mc instead of Ma because both showed similar correlation coefficients (table II). Both sexes showed similar amounts of Mc until the age of 12 except for girls who had greater ACG at 11 yrs and lower TCG at 10 yrs. Thereafter, boys had significantly higher mean Mc values than girls at each age group (table II), however, after accounting for height these differences were noticed only after 14 yrs (*P*<0.05).

**Table II.** Total and regional skeletal muscle mass (SMM) and fat free mass (FFM) variables by age and sex groups.

| Age | N | Sex | FFM (%) | FFM (kg) | SMM (%) | SMM (kg) | ACG (cm) | TCG (cm) | CCG (cm) |
| --- | --- | --- | --- | --- | --- | --- | --- | --- | --- |
| 8 | 25 | Girls | 77.5 ± 7.4 | 23.0 ± 3.2 | 30.1 ± 3.5 | 8.9 ± 1.3 | 16.6 ± 1.5 | 32.2 ± 2.4 | 23.4 ± 1.5 |
| | 46 | Boys | 85.7 ± 6.3 | 24.7 ± 2.9 | 40.3 ± 3.5 | 11.6 ± 1.4 | 16.5 ± 1.3 | 32.7 ± 3.0 | 23.1 ± 1.5 |
| | 71 | All | 82.8 ± 7.8 | 24.1 ± 3.1 | 36.7 ± 6.0 | 10.7 ± 1.9 | 16.5 ± 1.4 | 32.5 ± 2.8 | 23.2 ± 1.5 |
| 9 | 31 | Girls | 73.8 ± 6.1 | 25.7 ± 2.2 | 28.4 ± 2.8 | 9.9 ± 1.2 | 17.0 ± 1.2 | 33.3 ± 2.3 | 23.6 ± 1.5 |
| | 54 | Boys | 82.9 ± 7.5 | 27.9 ± 3.8 | 38.6 ± 3.2 | 13.0 ± 1.9 | 17.5 ± 1.8 | 34.3 ± 3.1 | 24.4 ± 2.0 |
| | 85 | All | 79.7 ± 8.3 | 27.1 ± 3.5 | 35.0 ± 5.8 | 11.9 ± 2.2 | 17.3 ± 1.6 | 33.9 ± 2.9 | 24.1 ± 1.9 |
| 10 | 24 | Girls | 77.4 ± 5.6 | 28.4 ± 5.5 | 30.4 ± 2.4 | 11.2 ± 2.5 | 17.1 ± 2.1 | 34.7 ± 3.4 | 25.5 ± 2.5 |
| | 65 | Boys | 80.4 ± 7.2 | 29.0 ± 3.9 | 39.7 ± 3.4 | 14.3 ± 2.0 | 17.6 ± 2.1 | 36.5 ± 2.9 | 25.3 ± 2.0 |
| | 89 | All | 79.6 ± 6.9 | 28.9 ± 4.4 | 37.2 ± 5.2 | 13.5 ± 2.5 | 17.5 ± 2.1 | 36.0 ± 3.1 | 25.3 ± 2.2 |
| 11 | 45 | Girls | 74.3 ± 6.7 | 32.5 ± 6.4 | 30.6 ± 2.8 | 13.4 ± 3.3 | 18.8 ± 2.4 | 37.4 ± 4.3 | 26.6 ± 2.7 |
| | 100 | Boys | 80.6 ± 6.8 | 31.8 ± 3.8 | 38.9 ± 2.9 | 15.3 ± 1.9 | 17.9 ± 1.7 | 37.5 ± 2.9 | 25.9 ± 1.9 |
| | 145 | All | 78.6 ± 7.3 | 32.0 ± 4.8 | 36.3 ± 4.8 | 14.8 ± 2.5 | 18.2 ± 1.9 | 37.4 ± 3.4 | 26.1 ± 2.2 |
| 12 | 38 | Girls | 76.2 ± 8.3 | 34.5 ± 4.1 | 31.6 ± 2.7 | 14.4 ± 2.6 | 19.1 ± 1.8 | 38.6 ± 3.4 | 27.5 ± 1.7 |
| | 114 | Boys | 79.5 ± 8.6 | 36.0 ± 5.7 | 38.5 ± 3.5 | 17.3 ± 2.6 | 19.2 ± 2.3 | 39.2 ± 3.3 | 27.6 ± 2.5 |
| | 152 | All | 78.7 ± 8.6 | 35.6 ± 5.4 | 36.8 ± 4.5 | 16.6 ± 2.8 | 19.2 ± 2.2 | 39.1 ± 3.3 | 27.6 ± 2.3 |
| 13 | 73 | Girls | 76.6 ± 5.9 | 38.3 ± 5.1 | 31.1 ± 2.5 | 15.6 ± 2.8 | 19.8 ± 1.8 | 39.2 ± 3.8 | 28.2 ± 2.0 |
| | 108 | Boys | 82.2 ± 8.9 | 41.1 ± 6.7 | 39.7 ± 3.8 | 19.9 ± 3.4 | 20.5 ± 2.5 | 41.5 ± 3.7 | 29.0 ± 2.8 |
| | 181 | All | 79.9 ± 8.3 | 40.0 ± 6.3 | 36.2 ± 5.4 | 18.2 ± 3.8 | 20.2 ± 2.3 | 40.6 ± 3.9 | 28.7 ± 2.6 |
| 14 | 73 | Girls | 74.3 ± 7.3 | 39.1 ± 4.5 | 31.0 ± 2.8 | 16.2 ± 2.4 | 20.1 ± 2.1 | 39.9 ± 3.4 | 28.5 ± 2.0 |
| | 119 | Boys | 82.2 ± 10.2 | 46.8 ± 8.0 | 39.8 ± 3.3 | 22.8 ± 4.4 | 22.2 ± 3.0 | 44.0 ± 4.6 | 30.7 ± 2.8 |
| | 192 | All | 79.2 ± 9.9 | 43.9 ± 7.9 | 36.6 ± 5.3 | 20.4 ± 5.0 | 21.4 ± 2.9 | 42.5 ± 4.7 | 29.9 ± 2.7 |
| 15 | 67 | Girls | 71.9 ± 7.8 | 41.3 ± 4.5 | 30.9 ± 2.9 | 17.5 ± 2.4 | 20.9 ± 2.1 | 41.1 ± 3.3 | 29.4 ± 1.9 |
|  | 131 | Boys | 82.6 ± 7.9 | 54.4 ± 8.0 | 40.5 ± 3.5 | 26.7 ± 4.0 | 24.0 ± 2.6 | 47.3 ± 3.8 | 32.9 ± 2.5 |
|  | 198 | All | 79.0 ± 9.3 | 50.0 ± 9.4 | 37.3 ± 5.6 | 23.7 ± 5.6 | 23.0 ± 2.8 | 45.3 ± 4.7 | 31.7 ± 2.9 |
| 16 | 52 | Girls | 73.8 ± 7.7 | 41.5 ± 6.2 | 36.3 ± 4.5 | 20.2 ± 2.8 | 20.6 ± 2.4 | 40.9 ± 4.0 | 29.4 ± 2.9 |
|  | 116 | Boys | 84.4 ± 7.5 | 56.4 ± 8.7 | 43.9 ± 4.4 | 29.1 ± 3.8 | 24.8 ± 2.8 | 47.8 ± 4.4 | 33.6 ± 2.8 |
|  | 168 | All | 81.2 ± 9.0 | 51.8 ± 10.6 | 41.7 ± 5.7 | 26.4 ± 5.4 | 23.6 ± 3.3 | 45.6 ± 5.3 | 32.3 ± 3.4 |
| 17 | 35 | Girls | 74.3 ± 6.4 | 43.7 ± 5.3 | 37.3 ± 3.1 | 22.0 ± 2.1 | 20.9 ± 1.4 | 42.5 ± 3.0 | 30.6 ± 2.0 |
|  | 72 | Boys | 84.9 ± 6.2 | 59.4 ± 9.7 | 43.8 ± 3.2 | 30.5 ± 4.2 | 25.2 ± 2.3 | 48.4 ± 4.1 | 33.8 ± 2.9 |
|  | 107 | All | 81.3 ± 8.0 | 54.2 ± 11.3 | 41.6 ± 4.4 | 27.7 ± 5.4 | 23.8 ± 2.9 | 46.4 ± 4.7 | 32.7 ± 3.0 |
| 18 | 15 | Girls | 74.9 ± 6.9 | 42.8 ± 5.3 | 37.2 ± 3.6 | 21.3 ± 2.4 | 20.8 ± 2.1 | 42.1 ± 3.9 | 29.5 ± 2.4 |
|  | 35 | Boys | 87.6 ± 4.6 | 61.5 ± 6.5 | 45.0 ± 2.7 | 31.5 ± 2.8 | 25.8 ± 2.0 | 49.1 ± 2.6 | 34.8 ± 2.0 |
|  | 50 | All | 83.5 ± 8.1 | 55.4 ± 10.8 | 42.5 ± 4.7 | 28.1 ± 5.5 | 24.3 ± 3.1 | 46.8 ± 4.5 | 33.0 ± 3.3 |

FFM, fat free mass; SMM, skeletal muscle mass; ACG, arm corrected girth; TCG, thigh corrected girth; CCG, calf corrected girth. All, boys and girls combined.

### MO

Predicted age at PHV was 11-12 yrs in girls and 13-14 yrs in boys. At age of 11, 17.8 % of the girls were classified as post-PHV and 73.7 % at 12 yrs. In boys, 34.3 % were post-PHV at 13 yrs and 91.6 % at 14 yrs.

### Sport participation across age

More than two-thirds of participants (77.2 %) were involved in sports programs (n = 1081, 25.4 % girls, 38 missing). General linear model analysis revealed positive age x sport participation interaction for all muscle variables (F = 2.66 for SMM, *P*=0.003; F = 2.71 for ACG, *P*=0.003; F = 3,37 for TCG, *P*=0.000; F = 3.07 for CCG, *P*=0.001) but disappeared after accounting for height. But, after including sex as factor, the interaction age x sex x sport participation was significant for SMM (F = 2.53, *P*=0.005) and ACG (F = 2.10, *P*=0.022). Similar results were found with participants grouped by school-age periods for the interaction school-age x sex x sport participation (F = 4.37 for SMM, *P*=0.013; F = 4.14 for ACG, *P*=0.016).

The regression analysis of Mc and age showed parallel lines for all limb Mc variables across all groups split by sex and sport participation (SP boys, NSP boys, SP girls, and NSP girls), but the patterns varied after adjusting to height (figure 1). Moreover, comparisons between the slopes revealed faster muscle growth rate by year in active youth compared with the non-active group by sex for all variables (SP vs NSP: boys F= 11.59 and girls F= 12.09 for ACG; boys F = 11.91 and girls F=11.02 for TCG; boys F = 12.23 and girls F=11.26 for CCG; boys F = 11.92 and girls= 11.78 for SMM; F = 12.13 in both sexes for FFM; *P*≤0.001 for all slope comparisons).

**Figure 1.**
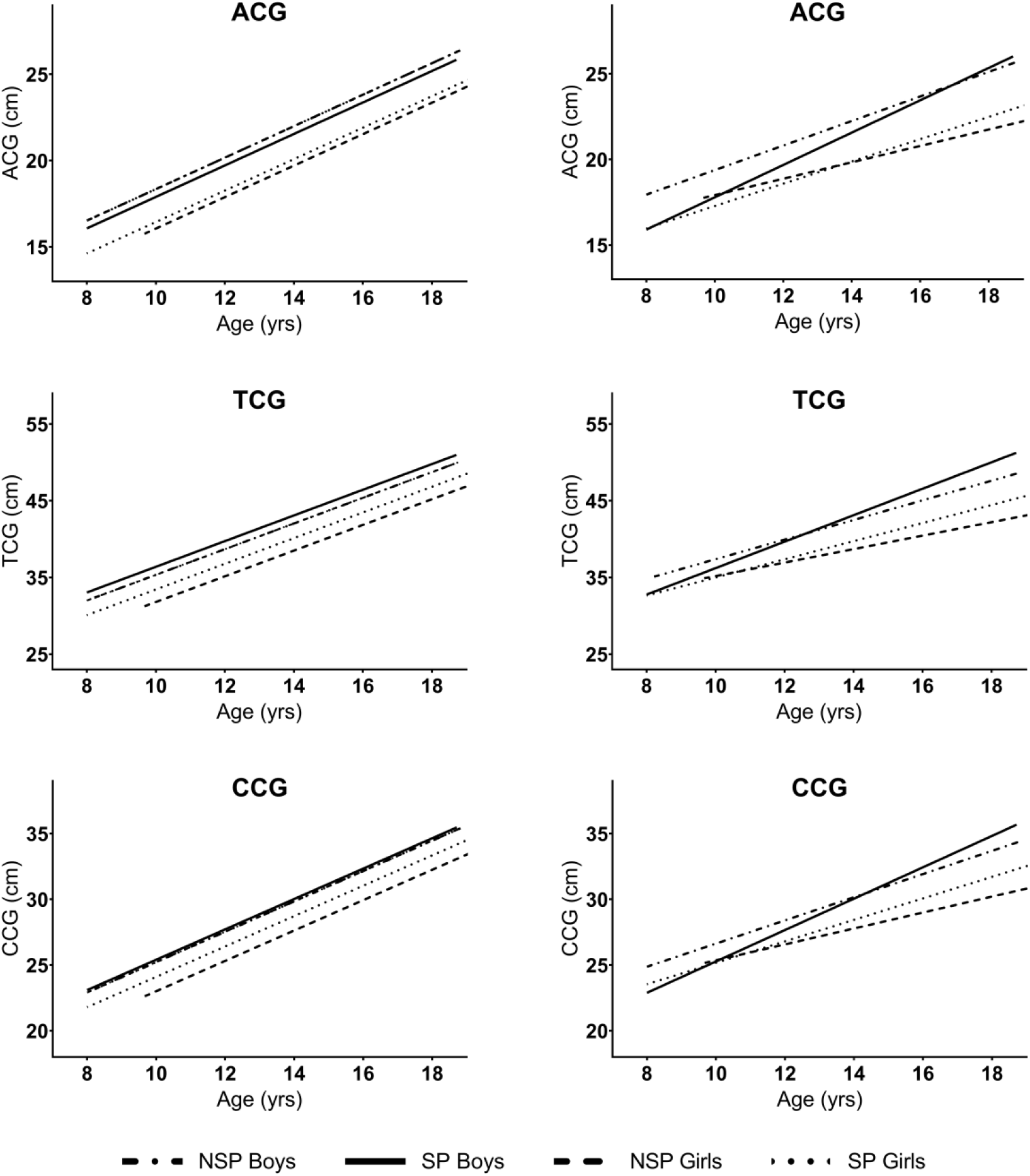
Regional muscularity evolution across age by sex and sport participation groups. Left panel: Regression analysis. Right panel: Regression analysis with height controlled for. SP, sport participant; NSP, non-sport participant. ACG, arm corrected girth; TCG, thigh corrected girth; CCG, calf corrected girth.

### SMM development along school-age periods

The analysis of SMM data showed increases in total and regional muscle along school-age periods in both sex and SP/NSP groups (*P*<0.05, figure 2). After adjusting these values for height and CA, only the active girls and boys increased their total SMM between all school periods, except between P-S to S-S. For the entire school period, SP girls and boys gained approximately 2 kg of SMM from P-S to H-S (2.18 kg for girls, 95% CI = 0.25 to 4.11; 2.15 kg for boys, 95% CI = 1.03 to 3.26). And from S-S to H-S, all groups showed total-body SMM gains except NSP boys (SP boys, 3.15 ±0.32 kg, 95% CI = 2.12 to 4.19; SP girls, 2.38 ±0.56 kg, 95% CI = 0.57 to 4.20; NSP girls, 2.75 ±0.62 kg, 95% CI = 0.74 to 4.77; all *P*≤0.01).

**Figure 2.**
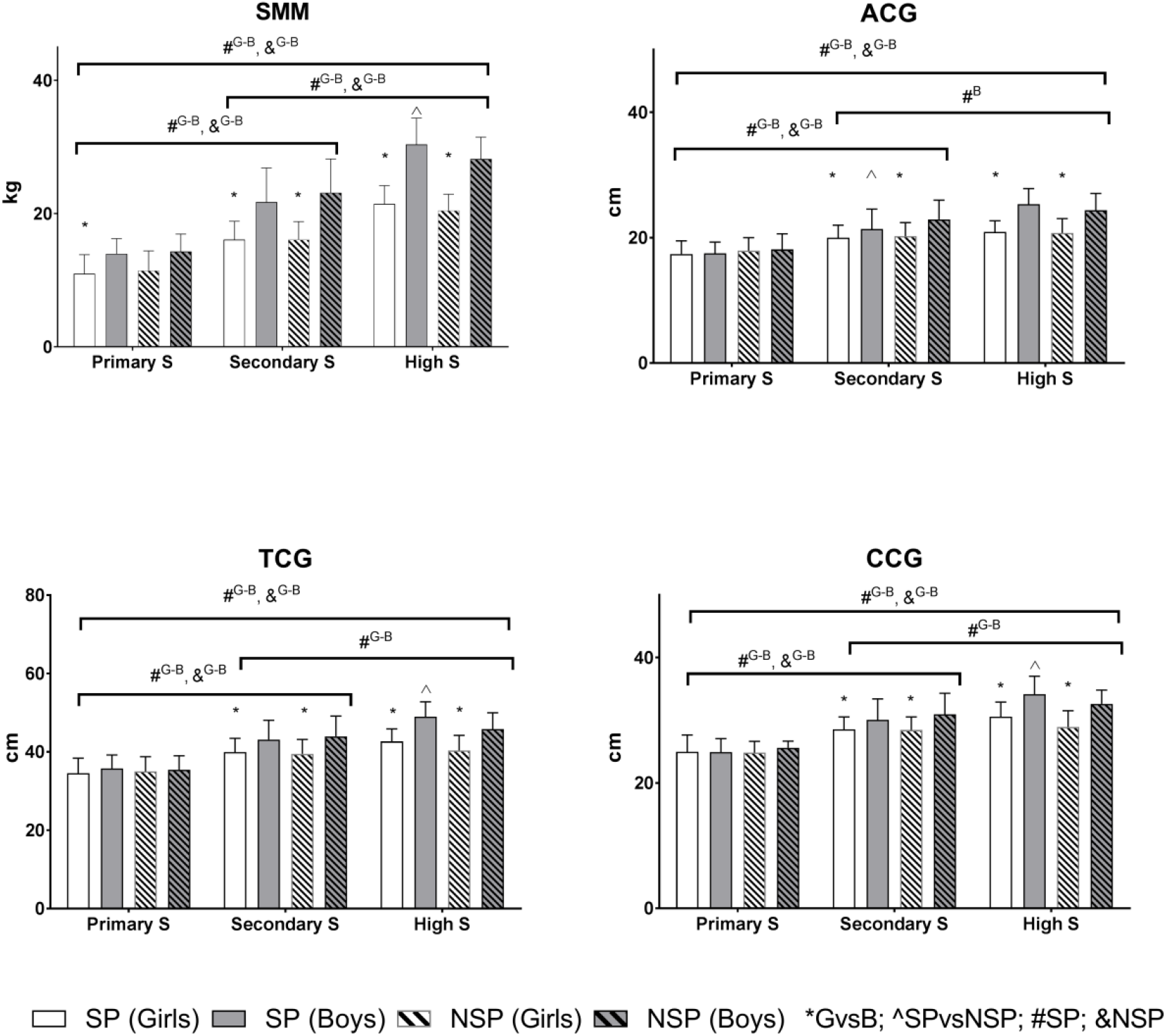
Comparison of total and regional muscularity across school-age periods. Symbols over brackets indicate *P* value <0.05 for comparisons *GvsB, girls and boys; ^SPvsNSP, sport participant and non-sport participant; #SP, same sex and sport participant; &NSP, same sex and non-sport participant; G, girls; B, boys; S, school.

Regarding regional muscularity, all groups increased Mc along school periods, except from S-S to H-S, when only the SP groups significantly increased their limb muscles excluding ACG of active girls (*P*>0.05, figure 2). Nevertheless, when Mc were adjusted for height and CA only active boys’ ACG increased significantly from P-S to H-S (1.29 ±0.31 cm, 95% CI = 0.29 to 2.29, *P*≤0.01) and from S-S to H-S (1.43 ±0.28 cm, 95% CI = 0.50 to 2.35, *P*<0.0001).

Multiple comparisons between boys and girls, and SP/NSP groups within school-age periods revealed that boys had always more total-body SMM than girls, except in P-S where NSP children did not differ (supplementary material). This pattern remained for regional variables but in P-S (figure 2). Additional contrasts between SP and NSP by sex revealed that only active boys in H-S had higher amounts of total-body and lower limb SMM than the non-active ones (mean difference = 2.18 kg for SMM, 95 % CI = 0.15 to 4.21, *P*<0.05; 3.16 cm for TCG, 95 % CI = 0.94 to 5.38, *P*<0.05; 1.56 cm for CCG, 95 % CI = 0.13 to 2.99, *P*≤0.001) (figure 2).

These previous results partially changed after adjusting values for body size (height) and CA, in summary, boys continued to have significantly higher SMM than girls regardless sport participation. Similar results were found at regional level for S-S and H-S although not in P-S, where only SP group differed (supplementary material).

### Allometric scaling for height

Ln-ln regression analysis was performed to scale muscle variables to height (Ht) and find specific allometric constants for sex, sport participation and maturity groups. The coefficients for SMM:Ht and FFM:Ht were 3.12 (SE ±0.44) and 2.81 (SE ±0.03) respectively for the combined sample. Boys showed higher ratio values than girls for FFM (2.82 ±0.03 vs 2.55 ±0.06, F = 15.84, *P*<0.0001) and the three regional variables (ACG:Ht, 1.38 ±0.03 vs 0.97 ±0.06, F = 34.72; TCG:Ht, 1.21 ±0.02 vs 1.05 ±0.05, F = 9.67; CCG:Ht, 1.21 ±0.02 vs 0.99 ±0.04, F = 17.77; all *P*<0.01). Conversely, girls showed higher ratio than boys for SMM:Ht (3.14 ±0.09 vs 2.95 ±0.04; F = 4.53, *P*=0.034). In the analysis of sport participation by sex with adjusted CA, significant differences in the slopes were found between SP and NSP boys for SMM:Ht (slope: 3.01 ±0.03 vs 3.33 ±0.1; F = 7.19, *P*=0.008) whilst no differences were seen among girls.

The analyses carried out between maturational age groups showed significant differences between the three slopes of the different phases of puberty for regional and total SMM variables in boys (ACG, F = 20.34; TCG, F = 7.82; CCG, F = 4.34; SMM, F = 29.86; all *P*≤0.01) and for total SMM (F = 4.17, *P*=0.02) but only ACG for regional variable in girls (F = 7.65, *P*<0.001; figure 3). Thus, significantly lower slope values were seen in the pre- and post-pubertal boys compared to the pubertal group for all muscle variables (*P*<0.03) and, between pre-pubertal compared to pubertal girls only for ACG and SMM (*P*<0.02). The highest unit of muscle per height (slope value) was found at pubertal ages in boys (ACG, 1.31 ±0.05; TCG, 1.16 ±0.04; CCG, 1.09 ±0.04; SMM, 2.92 ±0.07) whereas at post-pubertal ages in girls for all variables except for TCG in the pubertal group (ACG, 1.14 ±0.09; TCG, 1.06 ±0.08; CCG, 0.97 ±0.06; SMM, 2.81 ±0.15; figure 3).

**Figure 3.**
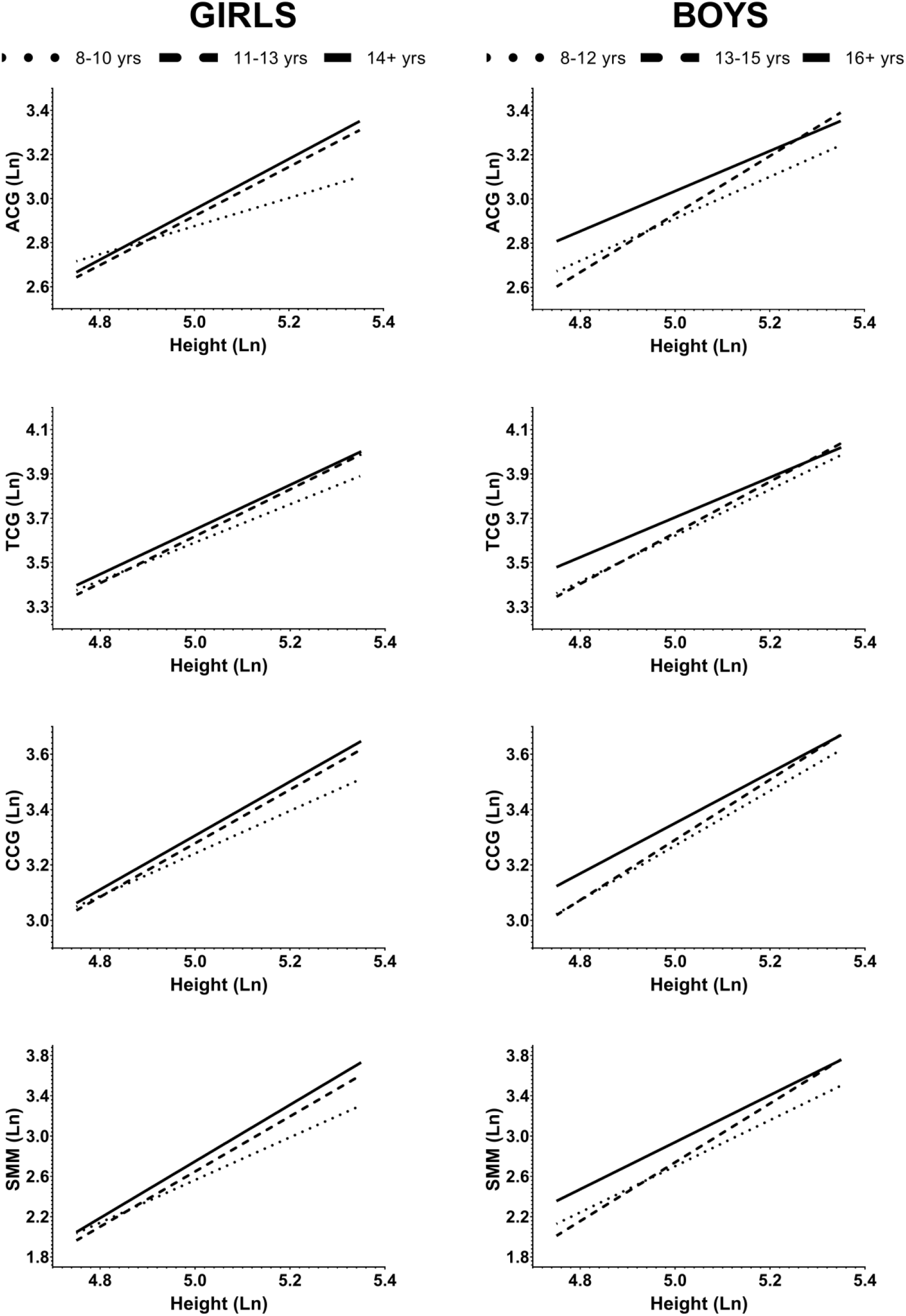
Ln-ln regression plots for total and regional muscularity with height by sex and maturation groups. ACG, arm corrected girth; TCG, thigh corrected girth; CCG, calf corrected girth; Ln, natural logarithm; yrs, years.

Relative to sport participation, the slopes between SP and NSP girls did not differ significantly for any muscle variable at any maturational age group (*P*>0.05). Only the Y-intercept value differed for TCG in post-pubertal girls (−1.41 ±0.42 for SP, and −1.03 ±0.66 for NSP; F = 5.67, *P*=0.018). For boys, only the pubertal group (13-15 yrs) showed significant differences between the slopes of SP and NSP for TCG (1.13 ±0.04 for SP, and 1.40 ±0.15 for NSP; F = 4.69, *P*=0.031). The Y-intercept of the regression differed between SP and NSP groups for ACG in pre- (*P*=0.001) and pubertal boys (*P*=0.005), and for SMM (*P*=0.012) only at pubertal ages. After excluding overweight and obese participants, differences between SP and NSP groups disappeared in boys.

## DISCUSSION

Data presented in this study showed trends for total-body SMM and FFM in children and adolescents across a wide-range of age. Regional indicators of muscularity based on anthropometric measurements were also reported for 13 specific age groups by sex. This simple and practical approach to quantify total and regional FFM can be useful for physicians, physical education teachers, and coaches working with pediatric and young populations. Our results may be helpful to inform of normal ranges of FFM, although the novel contribution of our study was the large dataset of scaling muscle-to-height and the role of sport participation and maturation in late childhood and adolescence.

Our observations of a non-linear pattern of FFM and SMM evolution with age and sex-related changes are consistent with the literature ^9,10,14^. Peak weight gain in girls occurred prior to puberty (10-11 yrs) due to a major contribution of the FM component whilst the opposite was seen in boys after PHV (15-16 yrs). This earlier expansion of FM than FFM sustained the hypothesis of a close link between the development of a fat energy store and maturation ^31^. Peak SMM was attained at late ages in both sexes which is consistent with data obtained by BIA ^9,14^ but differed from DXA measures when compared to Spanish adolescents girls ^32^.

Previous studies with different body composition techniques have reported similar ages for SMM spurt in boys (14-15 yrs) but not in girls (15-16 yrs). A longitudinal study in Canadian adolescents showed the peak velocity of lean body mass from DXA at ∼12 yrs in girls and ∼14 yrs in boys ^33^ and fastest growth of SMM from BIA up to 15 yrs in boys and 13 yrs in girls ^20^. For FFM derived from 4C model, growth spurt appeared to occur at earlier ages for boys (13-14 yrs) and girls (12-13 yrs) ^9^. These few discrepancies between studies may be explained by differences in study design, age group categories, sample size, ethnicity, body composition methodology and components analyzed ^34^.

Relative to regional muscularity, ACG, TCG and CCG changed significantly every year before and after the growth spurt period in our sample (girls, 11-13 yrs; boys, 12-15 yrs). Overall, boys and girls have similar amounts of Mc until the age of 12 which may be related to our maturational data reporting earlier ages in girls. This is supported by findings of sex heterogeneity in arm muscle area after 12 yrs ^16^ and reported values of ∼1.2 times greater body circumferences in girls than in boys between 10-12 yrs ^35^. However, when accounting for height differences, sex-dimorphism appeared after 14 yrs showing a smoother development in girls compared to a more noticeable growth in boys. Other body composition components (intramuscular fat or bone mass), biological changes, and intense physical activity (sport participation) may contribute to the observed variability across sex and age groups.

### Body Composition

The importance of FM for a healthy and normal muscle mass size has been recently highlighted in adults due to the physiological relationship between these two compartments ^36^. To explore this hypothesis, we analyzed the effect of total and regional variables of FM as covariates in the prediction of regional muscularity. Our results showed that total-body %FM was significantly associated with the size of ACG and TCG (*P*<0.001) but not with CCG (*P*=0.185). However, the results obtained for specific associated-region skinfold thicknesses showed a significant effect at the three regional sites (ACG and TCG, *P*<0.001; CCG, *P*=0.045), which may suggest that calf muscles development is independent of total but not regional fat.

### Sport participation

Despite the cross-sectional design of this analysis, our findings of the independent relationship between sport participation and SMM accrual are partially in agreement with longitudinal studies with smaller samples ^20^, which was also affected by body size (height) during the adolescent growth ^21^. This hypothesis was tested in our study by three different approaches. First, the analysis of the evolution of muscularity with age within sex and SP/NSP groups suggested that sport participation does not increase the rate of lower limb muscle growth across age, although our active volunteers had greater amounts of muscularity. Nonetheless, after accounting for body size (height), muscle growth rate with age was significantly higher in active girls than in the inactive. These results suggest a plausible sexual phenotype/dysmorphism of regional SMM influenced by sport participation.

In the second approach, with participants grouped by school-age periods, our results suggested sport participation does not contribute to differences in muscularity at any school period, but it may affect total SMM accrual in boys and girls across the educational periods (from P-S to H-S) independent of body size and CA. However, after puberty, increases in total SMM of girls were not influenced by sport participation (from S-S to H-S). At regional level, only boys’ ACG seemed to be affected by sport participation along the entire educational period.

Finally, the third approach used a scaling analysis between muscle variables and height (due to its non-linear relationship) to explore differences by sex, maturation and sport participation groups; these results are presented in the following paragraphs (body size adjustment and maturation).

### Body size adjustment

The conventional ratio FFM:Ht may remove partially the effect of body size and permit to compare data among samples from different studies. However, allometry represents a more elegant and accurate (ratios assume zero intercept and this introduce a bias to analyze body composition data) ^37^ way to describe how muscle and height scale with one another. Our findings showed sex differences in the relationship between FFM and height similar to those reported for body cell mass ^38^, which may confer our data construct validity and so physiological relevance. Our results are consistent with another study that reported FFM differences in early age stages of the adolescence growth period and later at 16-18 yrs for males and females ^39^.

The present study includes the scaling analysis of regional variables which may thus help understanding and interpretation of potential differences in the muscular development at regional and total-body level. The results showed power values close to value of 1 unit after normalizing regional indicators by height, being in all cases higher for boys than for girls, which may support the hypothesis that males have more SMM not only because larger body size but for other biological reasons (hormonal; see next section).

### Maturation

Due to the limited validity of maturity prediction equations ^40^, predicted ages at PHV were presented for informational use to compare with other studies. The scaling relationship between muscle and height through maturational groups (phases of puberty) showed the highest unit of muscle per height (slope) at pubertal ages in boys and post-pubertal in girls, except for TCG:Ht in pubertal girls. It seems that the maximum muscle growth rate per height may be influenced not only by pubertal stages and sex (like PHV ages in boys and after PHV in girls) but by regional lean mass distribution.

Regarding sport participation, only the NSP group of pubertal boys was found to have more log units of muscle per cm of height than their SP peers (thigh level only) although differences were explained mainly by the presence of overweight and obese participants. From this analysis it could be interpreted that participation in sports may not influence muscle growth in relation to height at any maturational age group. We speculate that the effect of sport participation in boys could be blunted by sexual maturation related to the increased testosterone secretion throughout puberty ^41^. More studies exploring how muscle scale to height are needed to elucidate whether this relationship is affected by sport participation before, during, and after PHV.

In general, our findings are in line with longitudinal data on the importance of physical activity for an improved lean mass accrual during growth spurt but partially disagree with our data relative to limbs ^21^. Overall, these results seem to support our initial hypothesis suggesting that sport participation could positively influence skeletal muscle growth during the developmental period and the scaling relationship between muscle and height may be dependent on sex, maturity, and regional distribution.

One of the main limitation of the study was the use of the anthropometric method to estimate Mc at thigh and calf level; nevertheless, this must be a valid option to be used in clinical settings and several studies have shown construct validity of this method in children and adolescents ^17,42^; for example, the upper arm index has been classically addressed as one of the most health-related parameters used in nutrition ^27^.

In summary, the results of this study are unique in reporting a large dataset of total and regional muscle mass variables across sex and sport participation groups (SP/NSP). Additionally, the current report extends data from previous studies by using a simple and low-cost technique and adding unique groups with specific traits; so, muscle girths data presented here may contribute to a better understanding of the SP phenotype in total versus regional SMM development. Finally, muscle scaling to height highlighted and confirmed that the anthropometric method is useful to observe and assess differences between sex and SP/NSP groups independent of maturation and body size. Further longitudinal studies exploring regional muscularity and regular participation in sports are needed to corroborate our results and emphasize the physiological significance of these reference data.

## PERSPECTIVE

This study may contribute to a better understanding of sports practice phenotype in total versus regional muscle development and will be helpful for pediatricians and other professionals working with children and adolescents to inform of expected ranges of fat-free and skeletal muscle mass. Furthermore, this dataset provides novel references for regional muscularity in sport participants and non-participants which are less commonly reported in the literature. Altogether, these resources must be important in the assessment of effectiveness of nutrition, exercise, and therapeutic interventions in children and adolescents. Finally, the methodology used in this study guarantees an accessible and low-cost procedure with applications in the majority of fields where skeletal muscle mass is relevant (sports coaches, physical education teachers, and nutritionists).

## Supporting information

Supplemental Table III

## Data Availability

All data produced in the present study are available upon reasonable request to the authors.

## Acknowledgements

Funded by the Spanish Ministry of Economy and Competitiveness (Grant: DEP2011-30565). We express our gratitude to the participants and their families, schools, and sports clubs for their collaboration. The authors thank Prof. Dr. Robert M. Malina for his insightful comments and suggestions during manuscript preparation, particularly on maturity assessment and analysis.

