## Supplemental Table III for "Total and regional skeletal muscle mass references by sport participation and body size in youth"

**Table III. Total and regional muscularity across school-age periods by sex and SP/NSP.**

| | PRIMARY-SCHOOL | | | |
|---|---|---|---|---|
| | **SMM (kg)** | **ACG (cm)** | **TCG (cm)** | **CCG (cm)** |
| **SP Boys** | 21.51 (0.30) | 20.99 (0.269) | 42.77 (0.39) | 29.30 (0.26) |
| **SP Girls** | 15.58 (0.40) | 19.54 (0.38) | 39.06 (0.64) | 28.02 (0.39) |
| **SP B vs SP G** (95% CI) | 5.93 (0.42) (4.57 to 7.29)¥ | 1.45 (0.37) (0.24 to 2.66)^ | 3.708 (0.562) (1.87 to 5.55)¥ | 1.273 (0.366) (0.07 to 2.47)* |
| **NSP Boys** | 21.24 (0.83) | 21.31 (0.705) | 41.80 (1.08) | 29.64 (0.72) |
| **NSP Girls** | 15.40 (0.47) | 19.79 (0.448) | 38.86 (0.75) | 27.47 (0.45) |
| **NSP B vs NSP G** (95% CI) | 5.84 (1.22) (1.85 to 9.82)¥ | 1.52 (1.06) (-1.93 to 4.98) | 2.95 (1.65) (-2.47 to 8.37) | 2.18 (1.08) (-1.35 to 5.71) |
| | **SECONDARY-SCHOOL** | | | |
| | **SMM (kg)** | **ACG (cm)** | **TCG (cm)** | **CCG (cm)** |
| **SP Boys** | 20.50 (0.13) | 20.85 (0.12) | 42.09 (0.17) | 29.37 (0.11) |
| **SP Girls** | 15.37 (0.18) | 19.69 (0.17) | 39.271 (0.29) | 28.11 (0.18) |
| **SP B vs SP G** (95% CI) | 5.13 (0.34) (4.01 to 6.24)¥ | 1.16 (0.31) (0.16 to 2.16)^ | 2.82 (0.46) (1.30 to 4.36)¥ | 1.26 (0.30) (0.27 to 2.25)^ |
| **NSP Boys** | 20.93 (0.33) | 21.92 (0.29) | 41.92 (0.43) | 29.66 (0.29) |
| **NSP Girls** | 14.91 (0.21) | 19.69 (0.19) | 38.31 (0.32) | 27.69 (0.20) |
| **NSP B vs NSP G** (95% CI) | 6.02 (0.54) (4.26 to 7.79) ¥ | 2.23 (0.48) (0.68 to 3.79)¥ | 3.61 (0.73) (1.22 to 6.00)¥ | 1.98 (0.48) (0.42 to 3.53)^ |
| | **HIGH-SCHOOL** | | | |
| | **SMM (kg)** | **ACG (cm)** | **TCG (cm)** | **CCG (cm)** |
| **SP Boys** | 23.65 (0.33) | 22.27 (0.29) | 42.58 (0.42) | 30.27 (0.28) |
| **SP Girls** | 17.76 (0.42) | 19.19 (0.40) | 39.16 (0.66) | 28.086 (0.40) |
| **SP B vs SP G** (95% CI) | 5.90 (0.54) 4.13 to 7.66)¥ | 3.09 (0.48) (1.5 to 4.67)¥ | 3.43 (0.72) (1.05 to 5.79)¥ | 2.18 (0.47) (0.64 to 3.72)¥ |
| **NSP Boys** | 22.97 (0.45) | 21.80 (0.38) | 40.36 (0.59) | 29.45 (0.39) |
| **NSP Girls** | 17.66 (0.46) | 19.31 (0.43) | 37.52 (0.72) | 26.93 (0.43) |
| **NSP B vs NSP G** (95% CI) | 5.03 (0.71) (2.72 to 7.35)¥ | 2.49 (0.61) (0.50 to 4.49)^ | 2.84 (0.96) (-0.32 to 5.99) | 2.52 (0.628) (0.47 to 4.58)^ |

Estimated means (SEM) and mean differences (SED) adjusted for height and chronological age; SEM, standard error of measurement; SED, standard error of difference; CI, confidence intervals; SP, sport participant; NSP, non-sport participant; B, boys; G, girls; SMM, skeletal muscle mass; ACG, arm corrected girth; TCG, thigh corrected girth; CCG, calf corrected girth; *$P$<0.05; ^$P$≤0.01; ¥$P$≤0.001.
